# Use of a Single Ventilator to Support Multiple Patients: Modeling Tidal Volume Response to Heterogeneous Lung Mechanics

**DOI:** 10.1101/2020.04.07.20056671

**Authors:** Vitaly O. Kheyfets, Steven Lammers, Jennifer Wagner, Karsten Bartels, Bradford Smith

**Affiliations:** University of Colorado Denver | Anschutz Medical Campus, Department of Bioengineering, Aurora, CO 80045; University of Colorado School of Medicine, Anesthesiology, Psychiatry, Medicine, and Surgery, Aurora, CO 80045

**Author notes:** **Summary Conflict of Interest Statement:** The authors report no conflicts. **Funding Information:** NA. **Notation of prior abstract publication/presentation:** NA.

## Abstract

The COVID-19 pandemic is creating ventilator shortages in many countries that is sparking a conversation about placing multiple people on a single ventilator. However, on March 26^th^ the American College of Chest Physicians (CHEST), along with other leading medical organizations, released a joint statement warning clinicians that attempting this technique could lead to poor outcomes and high mortality. Nevertheless, several hospitals around the United States and abroad are turning to this technique out of desperation (e.g. New York), but little data exists to guide their approach. The overall objective of this study is to utilize a computational model of mechanically ventilated lungs to assess how patient-specific lung mechanics and ventilator settings impact lung tidal volume (Vt).

**Methods:** We developed a single compartment computational model of four patients connected to a shared ventilator and validated it against a similar experimental study. We used this model to evaluate how patient-specific lung compliance (C) and resistance (R) would impact Vt under 5 ventilator settings of pre-set PIP, PEEP, and I:E ratio (suggested by Farkas, J.D. MD as an approach by hospitals to manage multiple patients on a single ventilator).

**Results:** Our computational model predicts Vt within 10% of experimental measurements. Using this model to perform a parametric study, we provide proof-of-concept for an algorithm to better match patients in different hypothetical scenarios of a single ventilator shared by more than one patient.

**Conclusions:** Assigning patients to pre-set ventilators based on their lung mechanics could be used to overcome some of the legitimate concerns of placing multiple patients on a single ventilator. We emphasize that our results are currently based on a computational model that has not been validated against any pre-clinical/clinical data. Therefore, clinicians considering this approach should not look to our study as an exact estimate of predicted patient tidal volumes.

## 1. Introduction

According to the World Health Organization (WHO), as of March 28^th^, 2020, the novel coronavirus (SARS-CoV-2, causing the disease COVID-19) initiated in Wuhan, China, has now been detected in 202 countries with over half-a-million confirmed cases worldwide ^1^.

The sudden surge in patients flooding intensive care units (ICUs) around the country has created a scarcity of mechanical ventilators ^2,3^, which has caused some centers to consider dual-patient (and sometimes even quad-patient) ventilation during critical ventilator shortages ^3-6^. However, the American College of Chest Physicians (CHES) and other leading organizations issued a statement on March 26^th^, 2020, warning practitioners not to attempt this practice ^2^. They list numerous important safety concerns for ventilating multiple patients on a single ventilator and warn that it could lead to poor outcomes and increased mortality. They also correctly point out that previous citations experimenting with this technique have also cautioned against using it ^4,7,8^. Nevertheless, faced with few good options, New York-Presbyterian Hospital and Columbia University have distributed a protocol for this approach ^9^.

Ventilation of any patient must be done with great care to avoid ventilator induced lung injury (VILI) ^10^ that damages the lung through the combined effects of tissue overdistension (volutrauma) ^11-15^, cyclic derecruitment and recruitment of small airways and alveoli (atelectrauma) ^16-20^, and inflammatory effects (biotrauma) ^21-24^. This is particularly of concern when attempting to ventilate multiple patients on a single machine because ventilator adjustments are applied to both patients, making titration to avoid VILI challenging. In this study, we utilized a computational modeling approach to evaluate the efficacy and considerations of ventilating multiple patients on a single ventilator.

This manuscript has the following overall objective: *J. D. Farkas MD has suggested a setup of 5 ventilator configurations* ^*3*^, *which could each support 4 patients. A pre-set fraction of inspired oxygen (FiO*_*2*_*), positive end-expiratory pressure (PEEP), peak inspiratory pressure (PIP), and inspiratory:expiatory ratio (I:E) is suggested for each ventilator. Based on these 3 controlled parameters for each ventilator, we provide a graphical reference for choosing the correct ventilator for a simulated patient, based on lung compliance (C) and resistance (R), to achieve the desired tidal volume (Vt)*.

We will also provide a discussion of possible complications to be considered in a clinical setting when supporting more than one patient on a single mechanical ventilator.

## 2. Methods

### Model Development

We developed a computational single compartment lung model that provides an estimate of pressure (P) and volume (V) in the lungs for a given patient’s lung compliance (C [=] mL/cmH2O), resistance (R [=] cmH2O-s/L), and time-dependent ventilation pressure (P_vent_ (t) [=] cmH2O). The model of a single patient (Fig. 1a) was expanded to consider four patients (Fig. 1b) connected to one mechanical ventilator providing pressure-controlled ventilation. Modeling details are provided in Appendix A.

**Figure 1.**
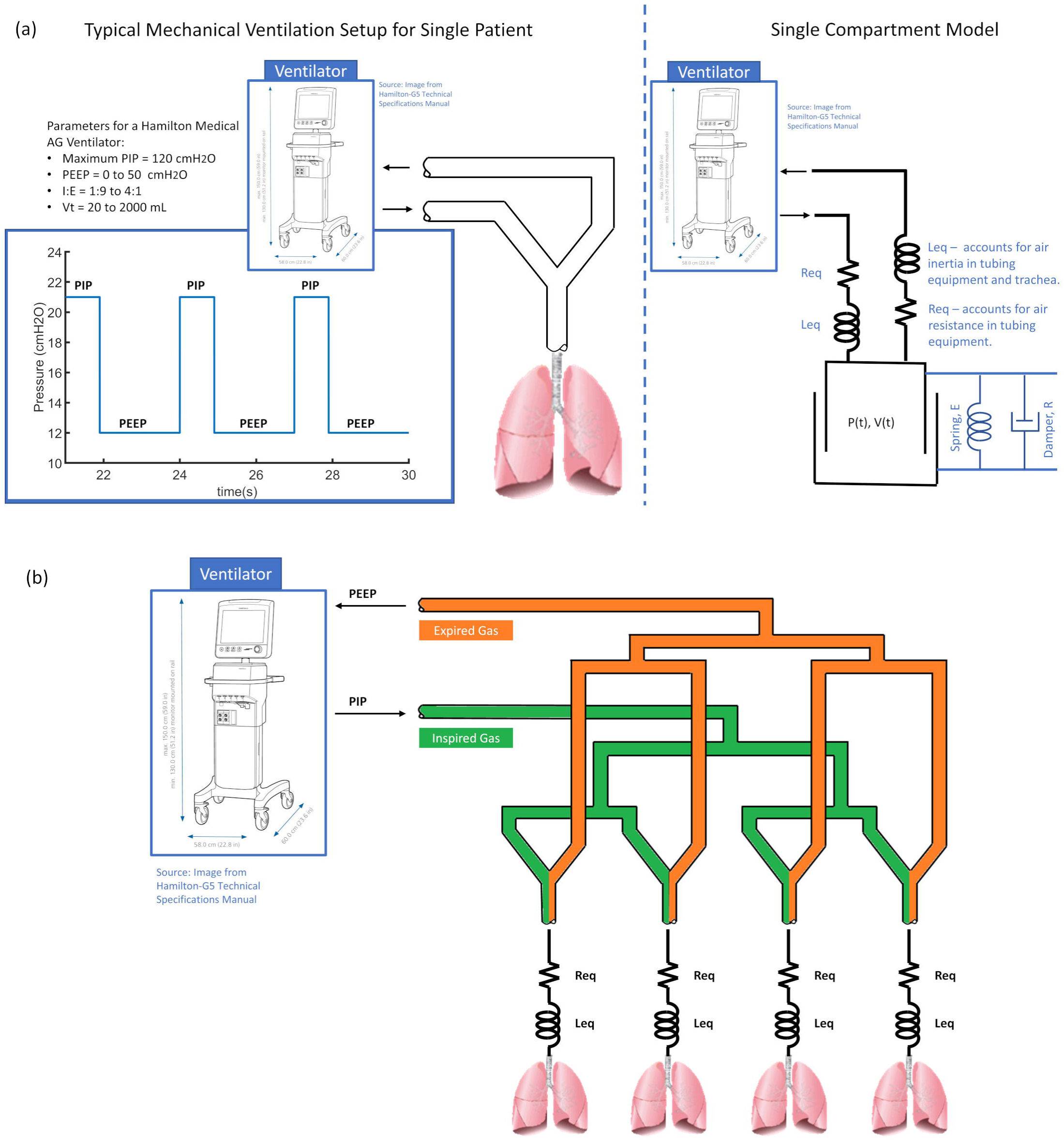
(a) Schematic showing single patient ventilation with corresponding single compartment model. (b) 4 patients on a shared single ventilator simulated using four single compartment model.

**Figure 1:**
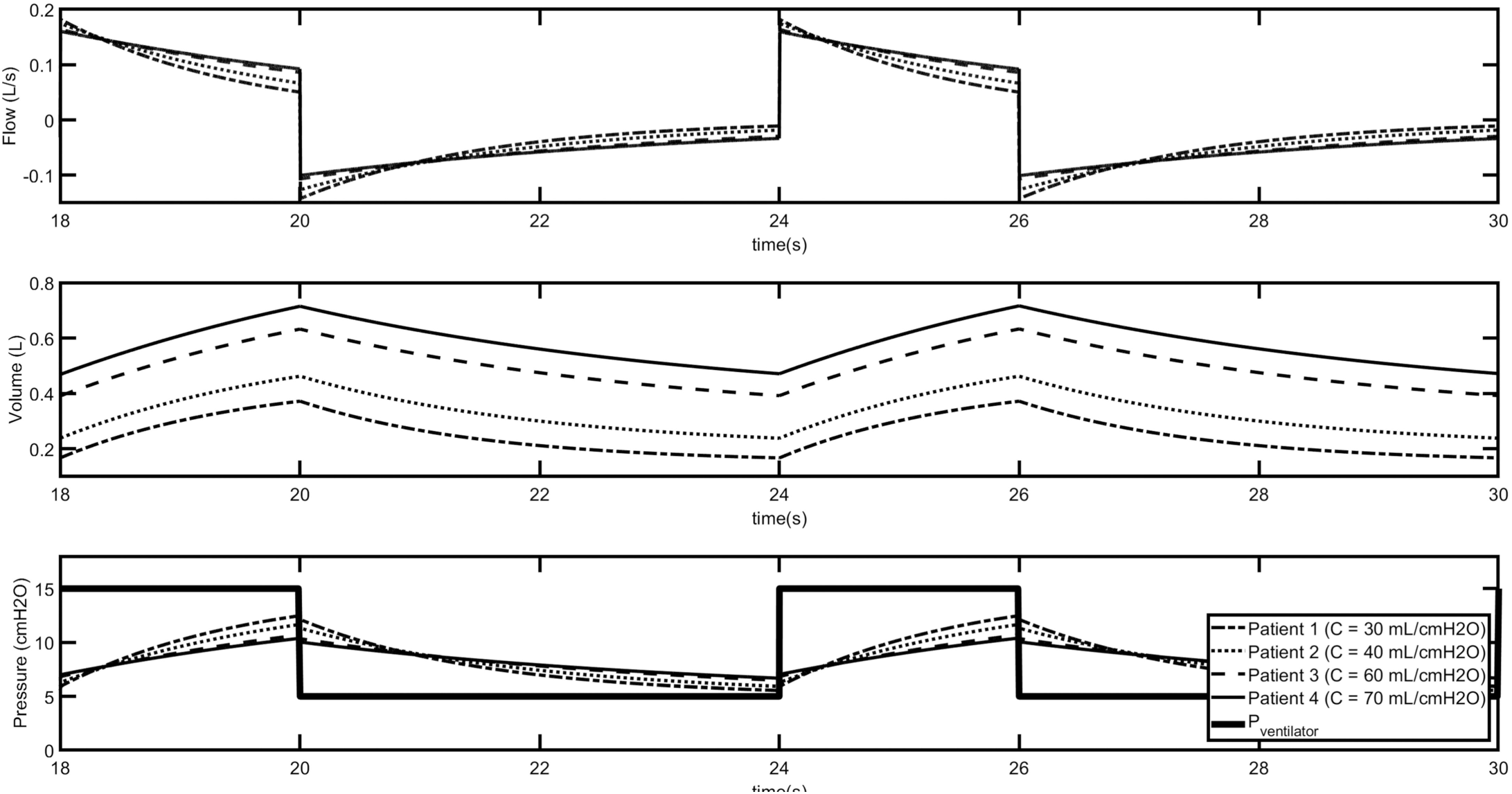
Simulated results for 4 patients at different values of lung compliance (C). Each patient was simulated using PIP = 15 cmH2O, PEEP = 5 cmH2O, lung resistance of 50 cmH2O-s/L, tubing resistance of 1.836 cmH2O-s/L, and tubing+airway inductance (Leq) of 0.02 cmH2O-s^2^/L.

The airflow provided to any patient is governed only by the pressure differential between the lung and the trachea. As such, during pressure-controlled ventilation, the lung mechanics of one patient should not significantly impact gas delivery to the other patients. However, there are certain theoretical scenarios that could present complications when simultaneously ventilating multiple patients (see Discussion).

### Model Validation

The model was validated against published experimental results in ^4^ and revealed limits of agreement for Vt -after correcting for bias-to be within 10%. The mean relative error in Vt between 16 model simulations and experiments was 2.13%. More validation details are outlined in Appendix B.

### Model Utilization

The model was used to perform a parametric study of R and C for the 5 ventilation scenarios proposed by J. D. Farkas ^3^ (see Table 1).

**Table 1.** Proposed constant ventilation settings ^3^ for continuously running ventilators modified to ventilate up to 4 patients.

|  | *PIP (cmH<sub>2</sub>O)* | *PEEP (cmH<sub>2</sub>O)* | *I:E* |
| --- | --- | --- | --- |
| ***Ventilator 1*** | 20 | 10 | 30:70 |
| ***Ventilator 2*** | 26 | 14 | 30:70 |
| ***Ventilator 3*** | 30 | 18 | 30:70 |
| ***Ventilator 4*** | 35 | 22 | 30:70 |
| ***Ventilator 5*** | 35 | 22 | 70:30 |

## 3. Results

Figure 2 shows simulation results for 4 patients with differing lung compliance values (C = 30,40,60,70 mL/cmH2O) on one ventilator. A constant resistance (R = 50 cmH2O-s/L) was used with ventilator settings: PIP =15 cmH2O; PEEP = 5 cmH2O; I:E = 30:70, respiratory rate (RR) = 20 breaths per minute. The tubing running from the ventilator to the patinets was considered to have a resistance (Req) of 1.836 cmH2O-s/L and inductance, Leq = 0.02 cmH2O-s^2^/L. The model accurately predicts expected trends in inspiratory/expiratory flow rate, tidal volumes and ventilator pressures. As expected, increasing compliance has a significant impact on decreasing tidal volume when holding ventilator pressure, resistance, and PEEP constant.

### Tidal volume (Vt) trends at different values of R and C, when assigning a patient to 1 of the 5 ventilator systems list in Table 1

**Error! Reference source not found**. shows a tidal volume sensitivity study for the 5 ventilator configurations proposed by J. D. Farkas MD ^3^ and listed in Table 1. In all simulations, RR = 20 breaths/minute and Leq = 0.02 cmH2O-s^2^/L. A contour represents tidal volume (Vt [=] mL), which was corrected by 19.6mL according to our validation analysis (see appendix B). In theory, these kinds of contour plots can be used to determine which patients would be most successfully grouped together on each ventilator configuration. If the airway resistance and compliance are measured prior to adding a given patient to a shared ventilator configuration, an estimated target tidal volume can be achieved (see Discussion).

## 4. Discussion

We created a computational model of multiple patients on one ventilator to provide insights on the application of this approach due to equipment shortages in disaster situations.

### Notes on Implementation

When sharing ventilators, the patients can be matched to the ventilator instead of adjusting the ventilator to match the patient. As suggested by Farkas ^3^, 5 classes of ventilators could be configured with different PIP, PEEP, I:E, and FiO_2_. Each of these ventilators is targeted towards a different degree of lung injury severity and could support between 1 and 4 patients. The challenge is then to decide which type of ventilation (numbered 1 through 5 – see Table 1 and Fig. 4) is best suited for a patient based on the desired Vt which may be 6mL/kg of ideal body weight ^25^ or higher to prevent hypercapnia in COVID-19 ARDS patients ^26^.

**Figure 4.**
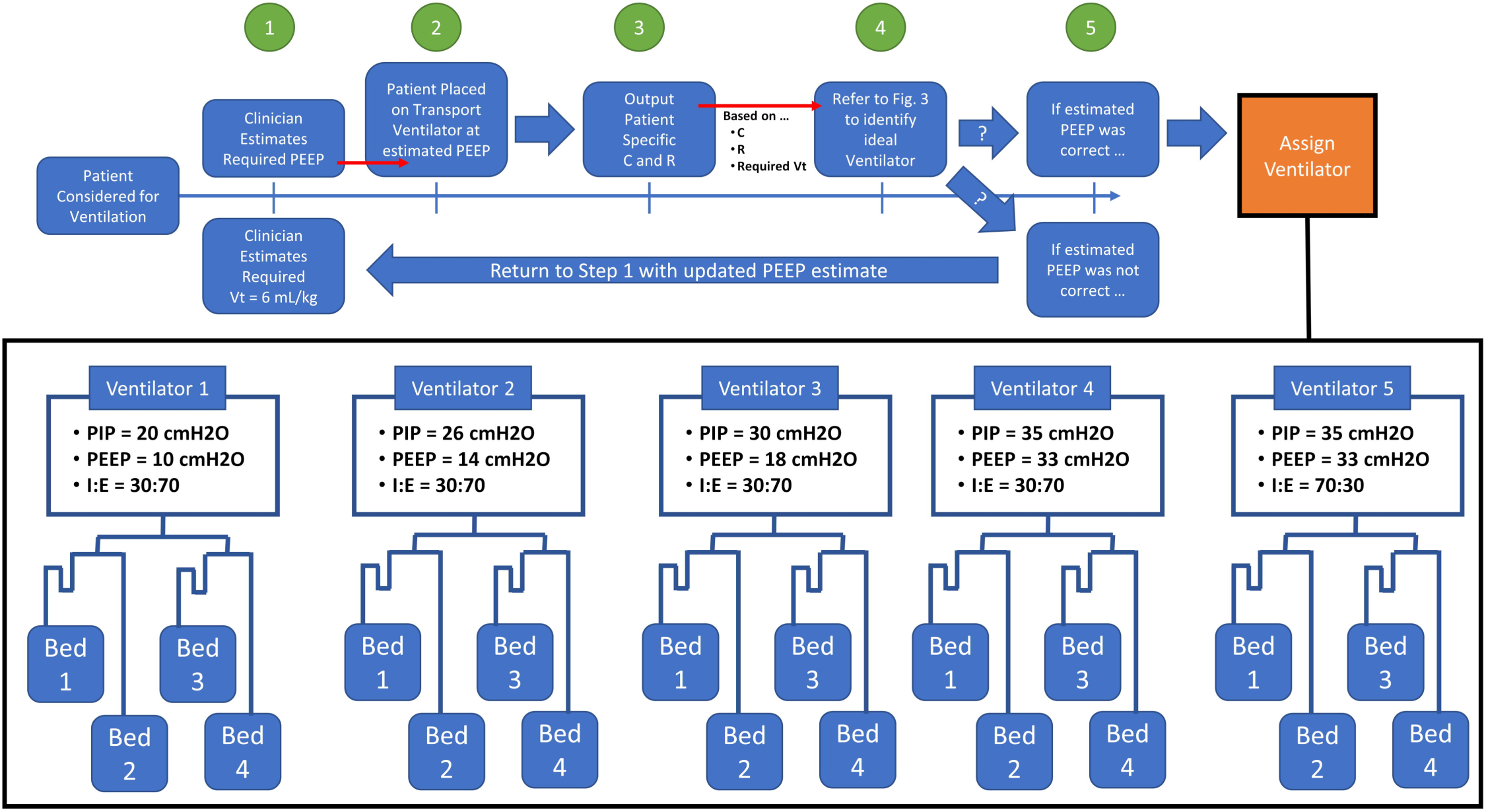
Potential clinical workflow for identifying the suitable ventilator based on patient tidal volume needs and lung mechanics (R, C). Note: Tubing length from the ventilator to all patients should be identical.

Our hypothetical clinical workflow would be initiated in a patient when 15 L/min O_2_ delivered via nasal cannula is insufficient to maintain oxygenation. These patients would be intubated and ventilated for 30-60 min using a transport ventilator that measures R and C while they are moved within the hospital. Based on those measurements, and the settings on the transport ventilator, the patient would be assigned to one of the five ventilator categories listed in Table 1 (or in Fig. 4). Alternatively, the patient could also be assigned to a single-patient ventilator if lung function is not compatible with shared ventilation. Patient lung mechanics (C and R) could be periodically re-evaluated, along with gas exchange, and if they improved, they could be moved to a lower-number ventilator (lower number ventilator indicated lower PIP, lower PEEP, and lower FiO2) ^3^. If the patient was deteriorating, the patient could also be moved to a higher-numbered ventilator. Ideally, a free ventilator would be kept near the shared ventilation ward to use for weaning and if lung function rapidly deteriorates in any patient. Of course, the operator would also confirm that the total required Vt for the 4 patients does not exceed the maximum available Vt from the ventilator.

#### Important Consideration

In many ARDS patients, lung compliance is non-linearly dependent on ventilation pressure due to volume-dependent tissue stiffness and pressure-dependent derecruitment ^27,28^. Therefore, the <u>value of C found on the transport ventilator is only valid for PEEPs similar to what is set on the transport ventilator</u>. To account for this, the transport ventilator used to measure the patient-specific C should be set at a PEEP the clinician judges to be within similar range of what they would expect for the PEEP value on the final long-term ventilator. For example, if the clinician anticipates that the patient will be placed on Ventilator 1 (PIP = 20 cmH2O, PEEP = 10 cmH2O, I:E = 30:70) then the transport ventilator PEEP should be set to approximately 10 cmH2O when measuring C. If the clinician reconsiders and decides to put the patient on Ventilator 4 (PIP = 30 cmH2O, PEEP = 22 cmH2O, I:E = 30:70) instead, C should be re-measured at a higher PEEP (e.g. between 20-24 cmH_2_O).

Figure 4 shows a hypothetical clinical workflow for selecting the proper ventilator using Fig. 3 based on patient lung mechanics and required tidal volume.

**Figure 3:**
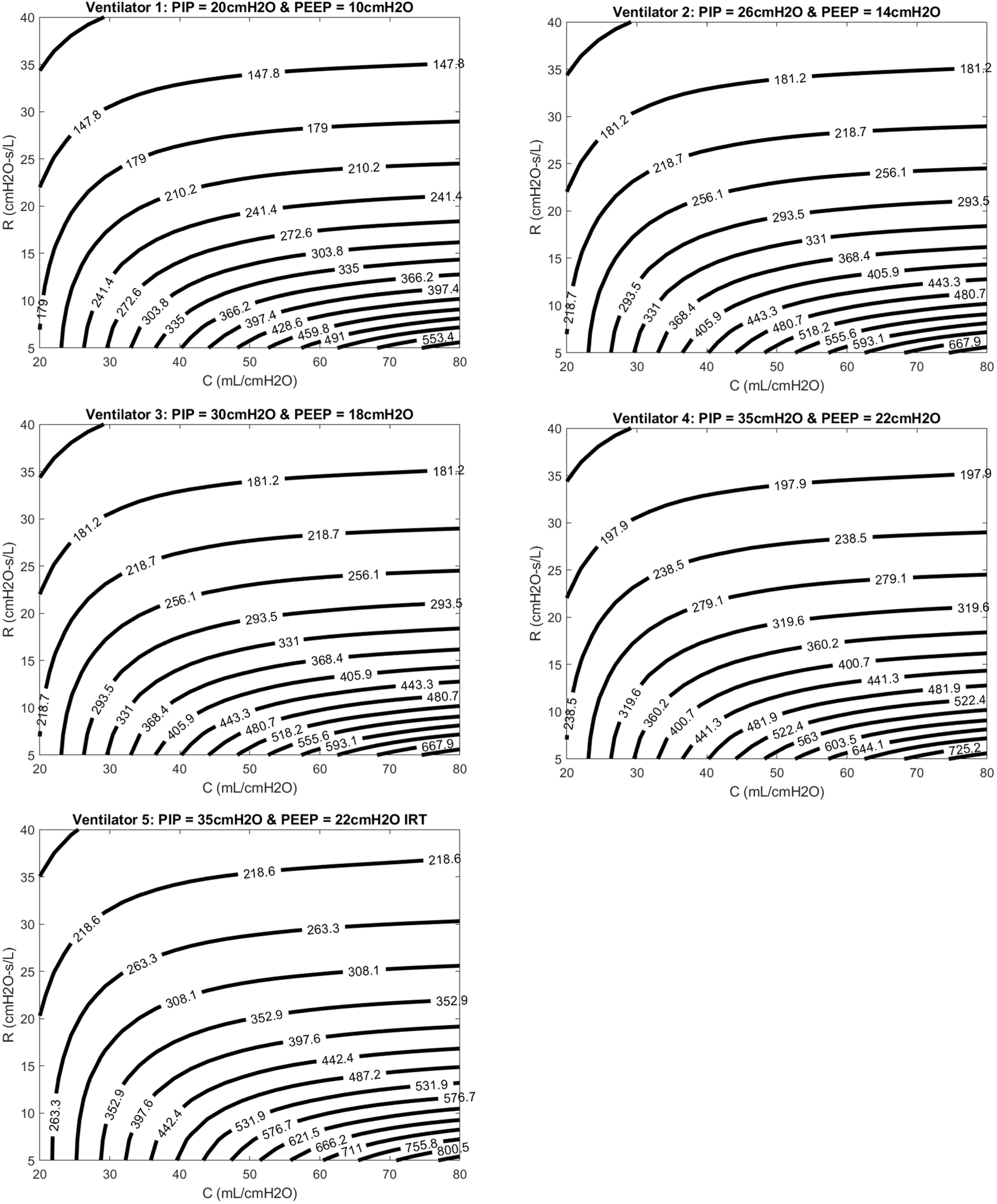
Contour plots showing ranges of estimated tidal volume for various R and C values (patient-specific lung mechanics), evaluated for 5 ventilator settings. Ventilator 5 was simulated with an inverse respiratory ratio: I:R = 70:30.

### Supporting experimental data

Current research on ventilating multiple patients using one machine includes using artificial lungs to measure flow distribution and tidal volume. The following review is not a comprehensive list of studies, but is focused on what we consider most relevant:

Branson *et al*. ^4^ performed an experiment using 2 dual-chamber test lungs to identify the impact of R and C on Vt, in a scenario where one machine would ventilate 4 lungs. This study was used to validate the current computational model. They found that differences in lung compliance, C, between patients has a higher impact of Vt inequality than resistance, R. Our model confirms this finding in patients with low compliance, but resistance becomes increasingly important for tidal volume in more compliant lungs due to the larger flow rates.

Paladino *et al*. ^5^ experimentally tested a four-limb ventilator circuit on 4 adult sized sheep, with a single ventilator. Although they appeared to have some issues initializing the experiment, all 4 sheep remained hemodynamically stable, successfully oxygenated, and ventilated for 12 hours.

Smith and Brown ^29^ successfully ventilated two human subjects on a single machine and found their combined tidal volumes to be 2 to 2.2 Liters. The subjects had different end tidal partial pressure of carbon dioxide (ETCO) at 10 minutes after the onset of ventilation (4.7 vs. 5.7 kPa). This offered proof-of-concept for clinical implementation, but suggests that additional fine-tuning is necessary for proper and safe ventilation.

### Consideration for ARDS patients and COVID-19

Unpublished technical notes from Italy have suggested two mechanical phenotypes for COVID-19 patients:

1. Good compliance – the patient is responsive to positive pressure. Lung injury can occur during expiration if PEEP is too high (causing overdistension).
2. Poor compliance – response should be similar to typical ARDS management.

The average compliance for a normal adult is 64 mL/cmH2O ^30^ and recent reports of COVID-19 ARDS lung compliance are in the range of 10 to 40 mL/cmH_2_O ^26,31^. Resistance varies between 8-12 cmH2O-s/L in normal subjects at 60 L/min of a square-wave inspiratory flow but could vary depending on endotracheal tube size and lung injury severity. However, both experimental studies by ^4^ and our simulation results (see Fig. 3) suggests that resistance is less important in patients with low compliance. It is important, however – as suggested by Fig 3-to account for resistance in patient with “good compliance.”

Recent studies from the Wuhan, China, population have also suggested that COVID-19 ARDS patients are not well responsive to high PEEP ventilation due to extremely high plateau pressures (> 45 cmH2O) ^31^. They propose a new mechanics-based index of lung recruitment that can be assessed by a single breath maneuver on any ventilator. This index could also be highly correlated with lung compliance and will be investigated in future studies.

### What could go wrong?

A joint statement by some of the leading respiratory organizations have strictly cautioned against using a single ventilator to support multiple patients ^2^ and list serious potential complications of this approach. Yet a webinar (https://register.gotowebinar.com/register/8748728666131694091) on 03/27/2020 informed listeners that sharing one ventilator between two patients is currently practiced at Columbia University College of Physicians & Surgeons New York-Presbyterian Hospital. Furthermore, they released a ventilator sharing protocol that could overcome some of these concerns ^9^. Our simulations may provide guidance to refining this protocol and further insights into some of these concerns (insight into those concerns provided by our simulations are provided in italics):

- Volumes would go to the most compliant lung segments. *During pressure-controlled ventilation, tidal volume will be based on compliance and driving pressure. It may be possible to match patients with similar lung mechanics to allow for appropriate tidal volumes*.
- PEEP, which is of critical importance in these patients, would be impossible to manage. *The ‘best’ PEEP for each patient will not likely be used. However, it remains to be seen if a ‘good’ PEEP for a set of similar patients provides acceptable outcomes*.
- Monitoring patients and measuring pulmonary mechanics would be challenging, if not impossible to measure.
- Alarm monitoring and management would not be feasible.
- Individual management for clinical improvement or deterioration would be impossible.
- In the case of a cardiac arrest, ventilation to all patients would need to be stopped to allow the change to bag ventilation without aerosolizing the virus and exposing healthcare workers. This circumstance also would alter breath delivery dynamics to the other patients.
- The added circuit volume defeats the operational self-test (the test fails). The clinician would be required to operate the ventilator without a successful test, adding to errors in the measurement.
- Additional external monitoring would be required. The ventilator monitors the average pressures and volumes.
- Even if all patients connected to a single ventilator have the same clinical features at initiation, they could deteriorate and recover at different rates, and distribution of gas to each patient would be unequal and unmonitored. The sickest patient would get the smallest tidal volume and the improving patient would get the largest tidal volume.
- The greatest risks occur with sudden deterioration of a single patient (e.g., pneumothorax, kinked endotracheal tube), with the balance of ventilation distributed to the other patients. *The simulations suggest that during pressure-controlled ventilation, which is the suggested modality* ^*9*^, *that changes in one patient do not impact gas delivery to the others. The tidal volume delivered to each patient is dictated by the difference between inspiratory and expiratory pressure, which is regulated by the ventilator. Of course, acute changes in one patient (e*.*g. pneumothorax, kinked endotracheal tube) could interfere with flow distributions to the other patients until the ventilator regulates itself back to the pre-set pressure waveform. This response should be investigated for different clinical ventilators in future experimental studies*.
- Finally, there are ethical issues. If the ventilator can be lifesaving for a single individual, using it on more than one patient at a time risks life-threatening treatment failure for all of them.

We also propose two additional factors that must be considered when placing multiple patients on a single ventilator:

- Although inertia is included in the current model, our set value for Leq is low enough that it does not significantly contribute to air flow dynamics during ventilation. Flow inertia plays an important role when measuring respiratory mechanics in intubated infants, which include higher ventilation frequencies and smaller tracheal tubes ^32^. In low frequency ventilation of adults, this is expected to have a minimal impact on gas delivery. However, our model does reveal that -if Leq is large enough (∼2 cmH2O-s^2^/L)-there could be a scenario where the tracheal pressure is lower than PEEP in a single patient during expiration. Therefore, the expired air from one patient, flowing through the expiratory tubing back to the ventilator, could theoretically flow into another patient. Again, the model would suggest that this possibility is in cases of high flow inertia.
- It is critical that the sum of Vt to all 4 patients not exceed the tidal capacity of the ventilator. For example, the experiment performed by Smith and Brown ^29^ to only two patients would have exceeded the 2L tidal capacity of the Hamilton-G5 SW 2.8x (Hamilton Medical AG, Switzerland) ventilator.

### Limitations

The serious clinical limitations of utilizing a single ventilator for 4 patients has been extensively outlined in previous clinical and experimental studies ^2,4,7,8^. Therefore, this manuscript will focus on the present limitations of our study.

Our overall objective is to provide a proof-of-concept graphic reference (see Fig. 3) for choosing the proper ventilator (or ventilator settings) using a computational model for a hypothetical patient with known lung compliance (C) and resistance (R), to achieve a desired Vt. In some clinics, acquiring patient-specific R and C values may not be possible. Furthermore, even in cases where patient specific R and C values are available, these values are likely to vary in a patient as his/her conditions improves or deteriorates. Therefore, it would have to be continuously re-evaluated based on clinical recommendations.

Some centers routinely change from supine to prone positioning during ventilation, which could drastically change the patient’s R and C values. In fact, recent studies with an admittedly small sample size have shown anecdotal data that alternating body positioning improved recruitability in COVID-19 patients ^31^. So -if additional studies support this finding-routinely needing to change body position could complicate the clinical workflow being discussed in this manuscript and proposed by ^3^. This could require the patient to be on a different ventilator when flipped from a supine to a prone position, which may not be logistically feasible when caring for a surge of critically ill patients.

Figure 3 is derived from a purely mathematical model, which has only been validated against a single experimental study ^4^ that was performed using a single type of ventilator and artificial lungs. Experimental values obtained from that study were crudely found by digitizing the figures in the published manuscript. Furthermore, the study only offered 12 points of comparison, with 4 additional points recorded under identical experimental conditions.

Therefore, simulation results executed under the same conditions will be identical, but the experiment will include variability. However, without doing the experiment ourselves, it is difficult to know if that variability is a result of measurement noise or a true variation of dynamics that can’t be accounted for in our computational model. Therefore, additional validation experiments will need to be performed in *in vitro*, animal, and clinical studies.

Finally, we employ a single compartment model to represent the acutely injured lung. This simplification was done in order to allow comparison of results to the limited measurements of lung function that are available from clinical ventilators. Although this representation describes the important features of lung mechanics, namely the elastic and resistive properties, it does not account for volume-dependent stiffening at high lung volumes. It also does not account for the time- and pressure-dependence of alveolar derecruitment ^33^. Although these factors are not explicitly simulated, they are represented in lung compliance.

## 5. Conclusion

We present a computational modeling approach that could serve for rapidly evolving research on the feasibility of sharing one single ventilator between more than one patient. *In vitro* and *in vivo* experiments suggests that this may not be impossible, but great care must be taken to avoid VILI and other potentially catastrophic complications. We present a hypothetical graphical guide for the scenario where 5 ventilators are set at different PIP, PEEP, and I:E, and the patient is assigned to a ventilator based on patient specific R and C, and the desired Vt. Although, we believe our findings may be helpful to develop a better understanding of the limitations of the shared ventilator concept, we need to strongly caution against applying them to patient care at this time.

## Data Availability

We are happy to share data and code if requested.

## Bibliography

1. WHO. 2020.

2. Joint Statement on Multiple Patients per Ventilator [press release]. 2020.

3. Farkas JD. PulmCrit – Splitting ventilators to provide titrated support to a large group of patients. 2020; https://emcrit.org/pulmcrit/split-ventilators/.

4. Branson RD, Blakeman TC, Robinson BR, Johannigman JA. Use of a single ventilator to support 4 patients: laboratory evaluation of a limited concept. Respir Care. 2012;57(3):399–403.

5. Paladino L, Silverberg M, Charchaflieh JG, et al. Increasing ventilator surge capacity in disasters: ventilation of four adult-human-sized sheep on a single ventilator with a modified circuit. Resuscitation. 2008;77(1):121–126.

6. Neyman G, Irvin CB. A single ventilator for multiple simulated patients to meet disaster surge. Acad Emerg Med. 2006;13(11):1246–1249.

7. Branson RD, Rubinson L. One ventilator multiple patients—What the data really supports. Resuscitation. 2008;79(1):171–172.

8. Branson RD, Rubinson L. A single ventilator for multiple simulated patients to meet disaster surge. Acad Emerg Med. 2006;13(12):1352-1353; author reply 1353-1354.

9. Beitler JR, Kallet R, Kachmarek R, et al. Ventilator Sharing Protocol: Dual-Patient Ventilation with a Single Mechanical Ventilator for Use during Critical Ventilator Shortages Columbia University College of Physicians & Surgeons New York-Presbyterian Hospital 2020.

10. Slutsky AS, Ranieri VM. Ventilator-Induced Lung Injury. New England Journal of Medicine. 2013;369(22):2126–2136.

11. Webb HH, Tierney DF. Experimental Pulmonary-Edema Due to Intermittent Positive Pressure Ventilation with High Inflation Pressures. Protection by Positive End-Expiratory Pressure. Am Rev Respir Dis. 1974;110(5):556–565.

12. Kolobow T, Moretti MP, Fumagalli R, et al. Severe Impairment in Lung-Function Induced by High Peak Airway Pressure during Mechanical Ventilation - an Experimental-Study. Am Rev Respir Dis. 1987;135(2):312–315.

13. Dreyfuss D, Soler P, Basset G, Saumon G. High inflation pressure pulmonary edema. Respective effects of high airway pressure, high tidal volume, and positive end-expiratory pressure. Am Rev Respir Dis. 1988;137(5):1159–1164.

14. Hernandez LA, Peevy KJ, Moise AA, Parker JC. Chest Wall Restriction Limits High Airway Pressure-Induced Lung Injury in Young-Rabbits. J Appl Physiol. 1989;66(5):2364–2368.

15. Carlton DP, Cummings JJ, Scheerer RG, Poulain FR, Bland RD. Lung Overexpansion Increases Pulmonary Microvascular Protein Permeability in Young Lambs. J Appl Physiol. 1990;69(2):577–583.

16. Seah AS, Grant KA, Aliyeva M, Allen GB, Bates JHT. Quantifying the roles of tidal volume and PEEP in the pathogenesis of ventilator-induced lung injury. Ann Biomed Eng. 2011;39(5):1505–1516.

17. Smith BJ, Bates JHT. Assessing the Progression of Ventilator-Induced Lung Injury in Mice. IEEE Trans. Biomed. Eng. 2013;60(12):3449–3457.

18. Smith BJ, Grant KA, Bates JH. Linking the Development of Ventilator-Induced Lung Injury to Mechanical Function in the Lung. Ann Biomed Eng. 2013;41(3):527–536.

19. Muscedere JG, Mullen JBM, Gan K, Slutsky AS. Tidal Ventilation at Low Airway Pressures Can Augment Lung Injury. Am J Resp Crit Care. 1994;149(5):1327–1334.

20. Slutsky AS. Lung injury caused by mechanical ventilation. Chest. 1999;116(Suppl. 1):9S–15S.

21. dos Santos CC, Slutsky AS. The contribution of biophysical lung injury to the development of biotrauma. Annual review of physiology. 2006;68:585–618.

22. Tremblay LN, Slutsky AS. Ventilator-induced injury: From barotrauma to biotrauma. P Assoc Am Physician. 1998;110(6):482–488.

23. Halbertsma FJJ, Vaneker M, Scheffer GJ, van der Hoeven JG. Cytokines and biotrauma in ventilator-induced lung injury: a critical review of the literature. Netherlands Journal of Medicine. 2005;63(10):382–392.

24. Uhlig S, Ranieri M, Slutsky AS. Biotrauma hypothesis of ventilator-induced lung injury. Am J Resp Crit Care. 2004;169(2):314–315.

25. Ventilation with Lower Tidal Volumes as Compared with Traditional Tidal Volumes for Acute Lung Injury and the Acute Respiratory Distress Syndrome. New England Journal of Medicine. 2000;342(18):1301–1308.

26. Liu X, Liu X, Xu Y, et al. Ventilatory Ratio in Hypercapnic Mechanically Ventilated Patients with COVID-19 Associated ARDS. American journal of respiratory and critical care medicine. 2020.

27. Harris RS, Hess DR, Venegas JG. An objective analysis of the pressure-volume curve in the acute respiratory distress syndrome. Am J Respir Crit Care Med. 2000;161(2 Pt 1):432–439.

28. Venegas JG, Harris RS, Simon BA. A comprehensive equation for the pulmonary pressure-volume curve. J Appl Physiol. 1998;84(1):389–395.

29. Smith R, Brown JM. Simultaneous ventilation of two healthy subjects with a single ventilator. Resuscitation. 2009;80(9):1087.

30. Henderson WR, Chen L, Amato MBP, Brochard LJ. Fifty Years of Research in ARDS. Respiratory Mechanics in Acute Respiratory Distress Syndrome. American journal of respiratory and critical care medicine. 2017;196(7):822–833.

31. Pan C, Chen L, Lu C, et al. Lung Recruitability in SARS-CoV-2 Associated Acute Respiratory Distress Syndrome: A Single-center, Observational Study. American journal of respiratory and critical care medicine. 2020.

32. Lanteri CJ, Petak F, Gurrin L, Sly PD. Influence of inertance on respiratory mechanics measurements in mechanically ventilated puppies. Pediatr Pulmonol. 1999;28(2):130–138.

33. Bobhate P, Guo L, Jain S, et al. Cardiac catheterization in children with pulmonary hypertensive vascular disease. Pediatr Cardiol. 2015;36(4):873–879.

